# An image based approach for predicting the effects of endocrine disrupting chemicals on human health using deep learning

**DOI:** 10.1101/2020.08.05.20168419

**Authors:** Pantelis Karatzas, Yiannis Kiouvrekis, Petros Stefaneas, Haralambos Sarimveis

**Author notes:** Corresponding author, (Yiannis Kiouvrekis). Email addresses* (Pantelis Karatzas), (Petros Stefaneas), (Haralambos Sarimveis), (Yiannis Kiouvrekis).

## Abstract

In recent years, deep neural networks, especially those exhibiting synergistic properties, have been at the cutting edge of image processing, producing very good results. So far, they have been able to successfully address issues of classification and recognition of objects depicted on images. In this paper, a novel idea is presented, where images of chemical structures are used as input information in deep learning neural network architectures aiming at the generation of Quantitative Structure Activity Relationship (QSAR) models, i.e. models that predict properties, activities or adverse effects of chemicals. The proposed method was applied to a case study of particular interest, which is the prediction of endocrine disrupting potential of chemicals. Two different deep learning architectures were applied. The produced ImageNet model proved successful, in terms of accuracy, performance and robustness on training and validation sets. The new approach is proposed to the community as an alternative or complementary method to current practices in QSAR modelling, which can automate and improve the creation of predictive models.

## 1. Introduction

Recent years have seen considerable developments in the fields of machine learning and neural networks in particular (Abiodun et al., 2018). New deep learning architectures and methodologies allow for the use of big data, such as sound and images, for developing models, taking decisions and drawing conclusions. Deep neural networks technologies enable us to successfully address issues such as recognition, image categorization and recognition of objects depicted – in certain instances more efficiently than a human agent could (Voulodimos et al., 2018; Wang et al., 2020). Increased access to data combined with improved computing power have enabled researchers and programmers to find new applications for neural networks at a very rapid rate (Amodei et al.). Very recently, deep learning methodologies have been used successfully in various problems in life sciences like drug discovery and microscopic and medical scan analysis (Ramsundar et al., 2019).

Chemometrics is the science of extracting information from chemical systems by data driven methodologies. Machine learning methods have been employed extensively in this particular field, with neural networks playing a dominating role (Marini et al., 2008). Various attempts have been made at devising systems that can depict chemical structures (Guha & Willighagen, 2012); ‘SMILES’ is one of these systems of depiction which is commonly accepted (G & KH., 2018). The Simplified Molecular-Input Line-Entry System (SMILES) is a standard notation which takes the form of a linear string describing chemical structures using short ASCII strings. These descriptions are vital to modern chemical information systems, but they do not necessarily allow computational techniques to be directly applied to them. Specific software, like the Chemistry Development Kit (CDK) (Steinbeck et al., 2003) has been developed to calculate quantitative descriptors that can form standard training data sets for training machine learning models for predicting activities of molecules, known as Quantitative Structure Activity Relationshiop Models (QSAR) models.

Deep learning methodologies have not yet used extensively in the field of chemometrics and many applications focus on the analysis of spectroscopical data (Chatzidakis & Botton, 2019). Deep learning has also been used as an alternative to other machine learning methodologies using standard descriptors of chemicals (Simmons et al., 2008). In this paper we present a novel idea for the application of deep learning in the field of chemometrics, which is based on the structural representation of chemical compounds as images and use of only these images as input information in the training process. Many molecular processors can automatically transform SMILES strings into 2D depictions or 3D image representations (Weininger et al., 1989; Weininger, 1988). The method is demonstrated on a specific case study, which is the prediction of Relative Binding Affinity (RBA) of potential endocrine disrupting chemicals. The results are very promising, taking into account that the only input information to the produced deep learning models is the image representations of the molecule and there is no need for descriptor calculation and variable selection, which are time consuming preprocessing steps, often involving a trial and error computational process.

## 2. Materials and methods

The endocrine system plays a central role in regulating metabolism, development, reproduction and behavior in all vertebrates. The hypothesis advanced concerning the presence of endocrine disruptors (Colborn et al., 1993) has led to new studies expressing concerns about the effects of endocrine disruption on health and the environment (Myers et al., 2004). Studies incorporate findings and methodologies from different fields, including toxicology, endocrinology, developmental biology, molecular biology, ecology, behavioral biology and epidemiology (Myers et al., 2004). An endocrine disruptor is defined as “an exogenous chemical substance or mixture that alters the structure or function(s) of the endocrine system and causes adverse effects at the level of the organism, its progeny, populations, or subpopulations of organisms, based on scientific principles, data, weight-of-evidence, and the precautionary principle” (Zoeller et al., 2012). Data collected from ecological studies, animal models, clinical observation of human subjects and epidemiological studies indicate that endocrine disrupting chemicals pose a significant risk to wild life and human health (Street et al., 2018). It is therefore of particular importance to develop a data driven model that predicts the endocrine disrupting potential of chemical compounds, which is the objective of this study.

The data set consists of 1,459 chemical structures. Based on experimental data, they have been labeled with values concerning their Relative Binding Affinity on a logarithmized scale (logRBA). The data were gathered from the EADB dataset (Estrogenic Activity Database) (Shen et al., 2013a; Ng et al., 2014; Shen et al., 2013b). The data subset used involves only the endpoints for species (human) and for logRBA. Images of the chemicals were generated using the chemistry development kit (CDK) (Willighagen et al., 2017) and the indigo open source software (Indigo, 2020). An example is given in Fig. 1 which shows the images generated for estradiol with SMILES C[C@]12CC[C@H]3[C@@H](CCc4cc(O)ccc34)[C@@H]1CC[C@@H]2O and formula ”C18H24O2” by the two software platforms. The reason for generating images from two software sources was data set augmentation, since the starting dataset is relatively small for deep neural network training. However, this did not improve neural network learning but, on the contrary, it prevented convergence. The model presented in the results sections used only the CDK generated images. In order to proceed with the development of the models, we classified the chemical structures into 3 classes according to their experimental response. The first class comprises structures which have response values in the [−3.328, −0.26] range, the second class comprises response values in the [−0.259, 0.824] range and the third comprises values in the [0.826, 2.857] range. The classes have been encoded as a One hot vector.

**Figure 1:**
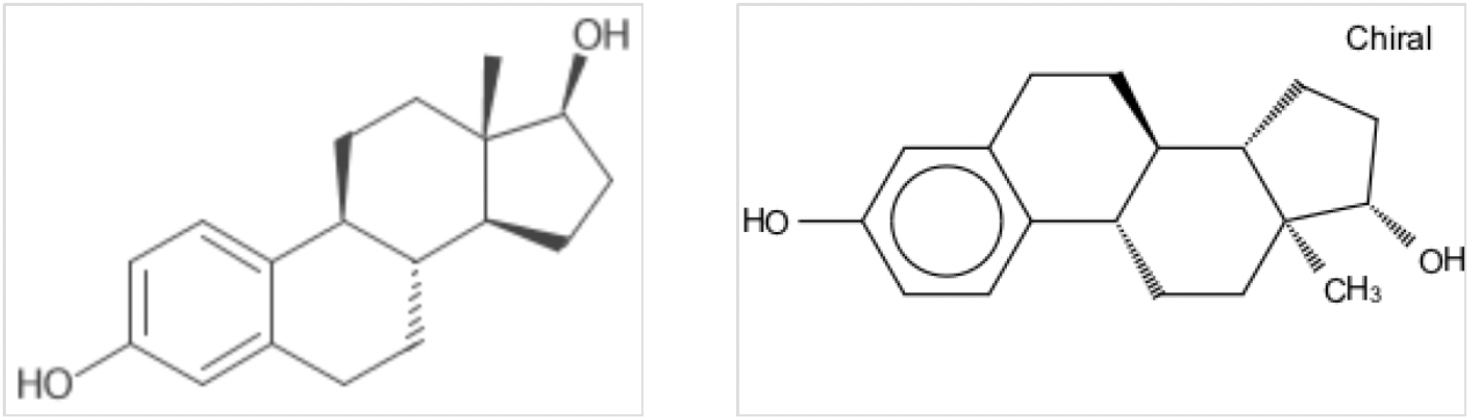
Two smiles images generated using two different software. Left: CDK produced SMILE image; Right: Indigo produced SMILES image.

The deep learning architecture employed for developing the models was AlexNet. AlexNet comprises eight layers; five of these are convolutional layers, some of which are followed by max pooling layers, and three are fully connected layers (Alom et al., 2019). AlexNet is quite similar to the older LeNet-5 architecture with two important differences: AlexNet contains more layers and employs the ReLU (Rectified Linear Unit) activation function that improves the training performance compared to the tanh or sigmoid activation functions used by LeNet-5. For the creation and the training of the models we used Tensorflow. The models have been trained on a pc with linux, an i3 cpu 16 gb of RAM and an nvidia GeForce 1070 gnu with 8 GB of RAM capable of running and training neural networks.

## 3. Results and discussion

The images were resized and pasted into white background so as to fit into squares as the input to a neural network. The dimensions used in modelling are 128 * 128, 200 * 200 and 256 * 256. 128 and 256 sized pictures favour the pooling layers of a deep neural network that can downsample the images to 8 * 8 sized convolutional kernels, or even 4*4. This way we can add many layers on the neural network if necessary. It should be noted that images have been normalized. Their pixel values were scaled so as to have a mean value of 0 and a standard Deviation of 1.

In order to proceed with training, the data set was split into random batches according to training needs. The memory capacity of the equipment available to the user plays a crucial role. In our case we conducted random sampling from batches of 42 images each. The batches were fed into the neural network to proceed with the training procedure. We proceeded employing two architectures of neural networks, namely the AlexNet type, as previously described, and in the second instance we used neural networks with Residual blocks.

The AlexNet network was constructed as follows: Three convolutional layers were used after the network input layer. Each convolutional layer was followed by a 2 * 2 pooling layer created by downsampling of the network input. Therefore, depending on the input, the final convolutional layer contained 16 * 16 filters times the filter depth for 128*128 sized images, 25 * 25 times the filter depth for 200*200 sized images and, in the case of 256 *256 sized images, 32 * 32 times the filter depth. The two final fully connected network layers consisted of 1024 nodes. The network output layer contained the three classes we need to predict. The activation function employed was ReLU, *f*(*x*)= *max*(0,*x*) as mentioned before.

We used two optimization functions, the classic Gradient Descent method and AdamOptimizer. The learning rate value was fixed to a relatively high value of 0.3, whereas typical learning rate values are in the range of 0.01 - 0.001. Lower values of learning rates resulted to slower convergence and smaller accuracy on the training data. On every network layer we used the dropout method to avoid over fitting the model. The input image size was not significant, since the accuracy of the model was affected by 1% at most, without exhibiting any patterns. The training accuracy graphs are presented below. The first graph (Figure 2) shows the evolution of accuracy during training. As anticipated, from a certain step and on wards (i.e. after step 5000) accuracy approaches 1:

**Figure 2:**
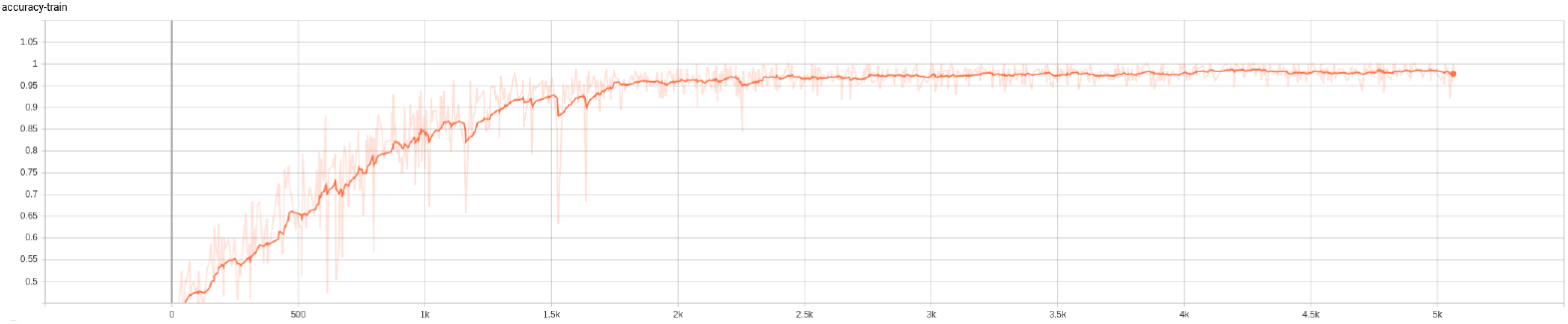
Evolution of accuracy on the training data for the AlexNet classification deep learning model.

Figure 3 is the corresponding accuracy graph for the validation data, where accuracy reaches a limit of 67%. Standard metrics in the field of machine learning were used for evaluating further the performance of the models. In particular, the confusion matrix also known as an “error matrix” (Weininger, 1988), refers to a certain matrix configuration which allows for visualizing the algorithm performance, typically in cases of supervised learning in classification models. Each row in the matrix represents occurrences in a predicted class, while each column represents occurrences in an actual class (or vice versa) (Voulodimos et al., 2018). It is named after the fact that it facilitates determining whether the system causes confusion in the various classes. The confusion matrix for the validation data set is shown in Table 1.

**Figure 3:**
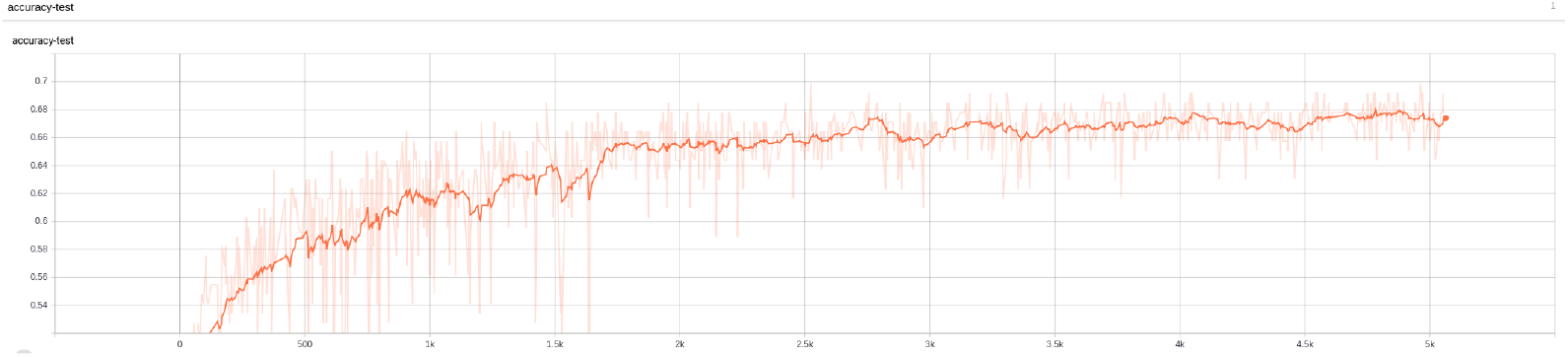
Evolution of accuracy on the test data for the AlexNet classification deep learning model.

**Table 1:** Confusion Matrix (absolute numbers and % accuracy on test images available in each class)

| n=146 | Number of images | Predicted class 0 | Predicted class 1 | Predicted class 2 |
| --- | --- | --- | --- | --- |
| Actual class 0 | 53 | 34 (64%) | 13 | 6 |
| Actual class 1 | 39 | 6 | 26 (66%) | 7 |
| Actual class 2 | 54 | 4 | 12 | 38 (70%) |

Matthews Correlation Coefficient (MCC), introduced by biochemist Brian W. Matthews in 1975, is used in machine learning to measure the quality of binary classifications (two categories). The coefficient takes into account both true and false positives and negatives and is generally considered as a balanced measure which can be employed even if the classes differ greatly in size. In essence, MCC is a correlation coefficient applied to observed and predicted binary classifications. It yields a value between -1 and 1. A coefficient of 1 represents a perfect prediction, 0 is better than a random prediction and -1 indicates complete disagreement between prediction and observation. The Matthews correlation coefficient has been generalized to the multi class problem, which is the case (Gorodkin, 2004) in our classification problem. Our value for the model’s predictions is MCC = 0.51.

For comparison purposes, we employed Residual nets as an alternative neural networks architecture, which has produced very good results in a number of modelling cases. Residual networks manages to go deeper than other networks with the utilisation of skip connections. This way they avoid the problem of the vanishing gradient by adding the activations of previous layers (He et al., 2016). They solve problem of CIFAR 10 (Krizhevsky, 2020) data set with an error rate of 4% (Angelov et al., 2016) and has good generalization performance for the data sets of PASCAL VOC 2007 and 2012 (Everingham et al., 2009) and COCO (Lin et al., 2014). We tried various levels in the network. The models created had either 10 or 15 residual blocks with 2 or 3 strides respectively followed by a fully connected layer with the outputs of the model. Again we used The Adam Optimizer with the value of 0.3. In Figures 4 and 5 the evolution of accuracy on the training and test sets is presented for a residual network of 10 residual blocks with a stride of 2 and a learning rate with the value of 0.3. The number of the parameters of the network proved computationally expensive. For the 8000 training steps we needed around 8 hours and thus we did not further proceed the training of the model.

**Figure 4:**
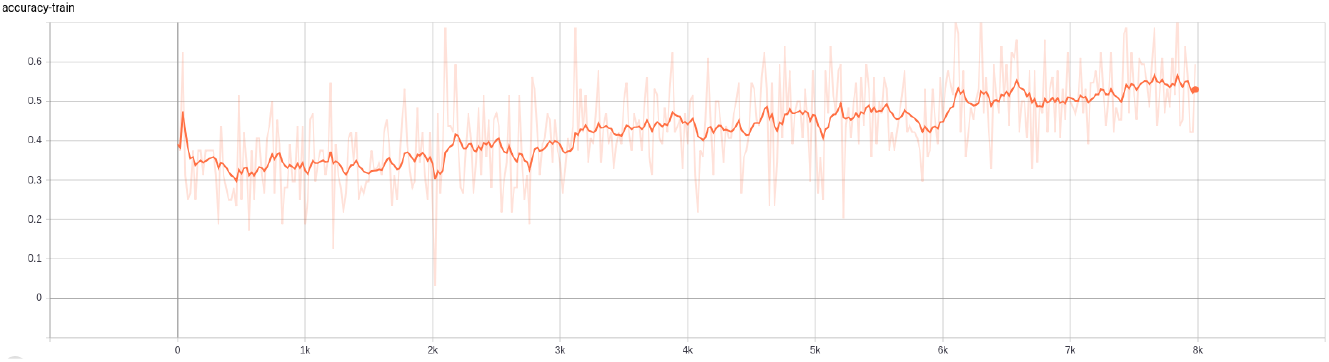
Evolution of accuracy on the training data for the Residual Net classification deep learning model.

**Figure 5:**
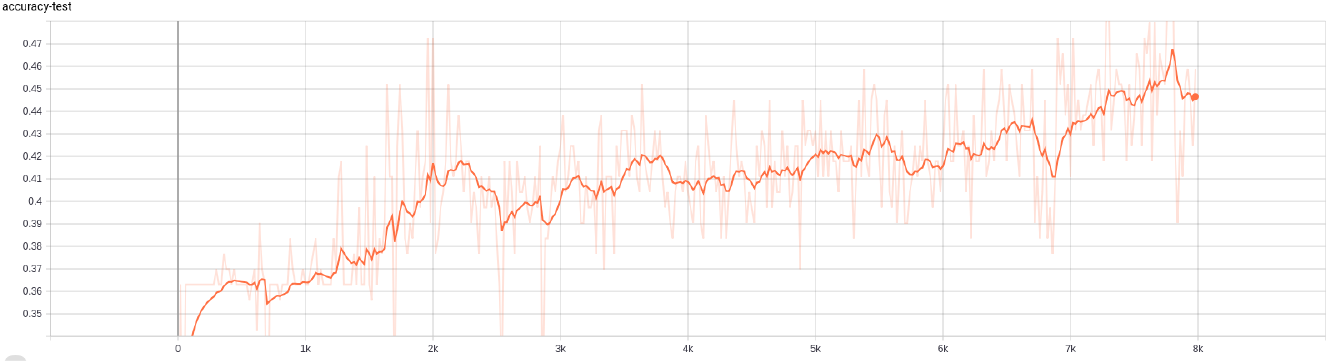
Evolution of accuracy on the test data for the Residual Net classification deep learning model.

We observe that the training procedure was not completed after 8000 iterations, which took more than 6 hours of computational time and the accuracy did not reach the level that was achieved using the AlexNet architecture. Therefore the AlexNet deep learning model was selected as the final classification model.

## 4. Conclusions

In this paper we demonstrated that 2D images of chemical structures could be the sole input information used in QSAR modelling, where deep learning architectures are employed. We investigated on the most adequate architecture that is capable of producing a reasonably accurate model. The accuracy and robustness of the model were evaluated on samples that were not used during the training procedure. The statistical metrics indicate that the proposed approach is very promising. It is of particular importance the fact that the proposed method does not require the calculation and selection of the most important molecular descriptors, which is the usual practice in QSAR modelling. Future research will focus on the use of 3D images, which will give additional details on the structures of chemicals. We will also investigate if combinations of images with standard calculated descriptors may further improve the results. Applications of the method on additional and possibly bigger data sets will further evaluate its performance, accuracy and robustness.

## Data Availability

The data that support the findings of this study are available from the first author, [PK], upon reasonable request.

